# Detecting heterogeneity of intervention effects using analysis and meta-analysis of differences in variance between arms of a trial

**DOI:** 10.1101/2020.03.07.20032516

**Authors:** Harriet L Mills, Julian PT Higgins, Richard W Morris, David Kessler, Jon Heron, Nicola Wiles, George Davey Smith, Kate Tilling

## Abstract

**Background:** Randomised controlled trials (RCTs) with continuous outcomes usually only examine mean differences in response between trial arms. If the intervention has heterogeneous effects, then outcome variances will also differ between arms. Power of an individual trial to assess heterogeneity is lower than the power to detect the same size of main effect.

**Methods:** Several methods for assessing differences in variance in trial arms were described and applied to a single trial with individual patient data (IPD) and to meta-analyses using summary data. Where IPD were available, regression-based methods were used to examine the effects of covariates on variation. An additional method to meta-analyse differences in variances with summary data was presented.

**Results:** In the single trial there was agreement between methods, and the difference in variance was largely due to differences in depression at baseline. In two meta-analyses, most individual trials did not show strong evidence of a difference in variance between arms, with wide confidence intervals. However, both meta-analyses showed evidence of greater variance in the control arm, and in one example this was perhaps because mean outcome in the control arm was higher.

**Conclusions:** Low power of individual trials to examine differences in variance can be overcome using meta-analysis. Evidence of differences in variance should be followed-up to identify potential effect modifiers and explore other possible causes such as varying compliance.

## 1. Introduction

In medical research we often examine the average effect of an intervention on a quantitative outcome by comparing mean differences between arms of a randomised controlled trial (RCT). However, individual responses to interventions may vary. For instance, the effectiveness of an intervention might decrease with age, or there might be subgroups for whom the intervention has no effect. In the era of personalised (or stratified) medicine, there is increasing interest in identifying these effect modifiers or subgroups ^1^.

We are focusing on trials with a continuous outcome, where the main effect is the mean difference between two arms of a trial. Identification of effect modifiers or subgroups is often approached by testing for statistical interactions. A potential effect modifier is specified (usually *a priori*, for RCTs) and the null hypothesis tested is that the effect of the intervention on the outcome does not vary over the levels of the modifier (i.e. that there is no additive interaction). However, a trial powered to detect such an interaction needs to be approximately four times the size of a trial powered to detect a similar magnitude of overall treatment effect ^2,3^. An even larger trial will be required if the subgroups are very different in size. Multiple testing can be a problem if interactions with many covariates are examined, with a risk of overfitting ^4^, although this can be avoided by using model selection methods ^5-8^. All these methods require knowledge of, and data on, the potential effect modifiers: if an effect modifier is not measured, then its interaction with the intervention cannot be tested. However, if there is effect modification, this should lead to a difference in variances between the intervention and control arms ^9-11^. Thus, an alternative way to investigate effect modification, without pre-specifying the effect modifiers, is to examine whether variance in the outcome differs between the arms of the trial ^9,12-14^. If variation is detected, this would then require further study to identify the effect modifiers, potentially needing individual participant data (IPD).

As with the test for a specific effect modifier, power to detect a difference in variances will be low in a single trial powered to detect a difference in means. However, just as meta-analysis of mean effects gives greater power to detect an average intervention effect, meta-analysis of differences in variance should give increased power to detect effect modification. A small number of meta-analyses in epidemiology and ecology have reported on differences in variance ^9,12,15-23^, with applications to RCTs and other types of comparative study. Most of them found evidence of a difference in variance between arms, with varying strength of evidence (eTable 1).

Here we describe and implement methods for examining the effect of an intervention on the variance of an outcome, both in a single trial (with individual participant data, IPD) and using meta-analysis to combine across trials (using summary data). We describe the assumptions behind each method, and we show how to conduct further analyses with IPD to investigate which variables might be causing the effect modification. We use simulations to show that decisions about when to examine the association between overall mean and variance should not be based on reported means and variances from individual trials and are only suitable for some types of outcome data. We then illustrate the methods applied to a single trial using data from an RCT of cognitive behavioural therapy (CBT) to treat depression, and to two meta-analyses based on summary data: one of RCTs using computer-based psychological treatments for depression, and one exploring the effect of statins on low-density lipoprotein (LDL) cholesterol.

## 2. Methods for examining difference in variance between trial arms

### 2.1 Examining differences in variance between two arms using data from one trial

We review methods briefly here, presenting more detail in Table 1 and formulae in eAppendix §2.

One approach is to test the null hypothesis of equality of variances between the arms, using Glejser’s ^24^, Levene’s ^25^ or Bartlett’s test ^26^; of these, only Bartlett’s can be calculated using summary data. A different approach is to estimate the difference in variances and its standard error, either using a linear model with non-constant variance (LMNCV), or directly using summary data, as we propose here. Finally, rather than a difference, the ratio of the standard deviations or the log of the variability ratio (logSDR, ^9,27^) can be estimated, together with their standard errors.

All methods and analyses were implemented in R (R Foundation for Statistical Computing, Vienna, Austria) and code is available online (https://github.com/harrietlmills/DetectingDifferencesInVariance).

**Table 1:** Methods for examining differences in variance between two arms, and for examining the relationship between mean and variation across the two arms. Further method details (and equations) are in the eAppendix §2. Code for each method in R is provided online (https://github.com/harrietlmills/DetectingDifferencesInVariance).

| Test name | Description | Minimum requirements and assumptions |
| --- | --- | --- |
| ***Testing differences in variance between two arms using data from one trial*** |  |  |
| Glejser's test (22) | The absolute values of the residuals from a standard linear model of outcome against treatment are regressed on the treatment indicator. | Requires IPD<br>Assumes normality<br>Can include covariates<br>Can be defined for k>2 arms |
| Levene's test (23) | Levene's test statistic has approximate F-distribution with 1 and $N - 2$ degrees of freedom | Requires IPD<br>Suitable for non-normal data<br>Can be defined for k>2 arms |
| Bartlett's test (24) | Bartlett's test statistic has approximate chi-squared distribution (1 degree of freedom) when variances are equal | Can be calculated using IPD or summary data (sample sizes, SD)<br>Assumes normality<br>Can be defined for k>2 arms |
| ***Estimating differences in variance between two arms using data from one trial*** |  |  |
| Linear model with non-constant variance (LMNCV) | A linear model that assumes a different residual variation in each arm. | Requires IPD<br>Assumes normality<br>Can include covariates<br>Can be defined for k>2 arms |
| Difference in variances (VD) | The difference in sample variances and its standard error are used to calculate a test statistic with an approximate normal distribution, so a t-test is used to compare variances. | Can be calculated using IPD or summary data (sample sizes, SDs)<br>Assumes normality |
| Ratio of variances (RoV), F-test | The ratio of sample variances between the two arms has approximate F-distribution with $N_0 - 1$ and $N_1 - 1$ degrees of freedom, if the true variances are equal. | Can be calculated using IPD or summary data (SDs)<br>Assumes normality |
| Log of the ratio of standard deviations (logSDR) (8, 25)<sup>a</sup> | The log of the ratio of standard deviations and the sampling variance are used to calculate a test statistic with approximate normal distribution, so a t-test is used to compare variances. | Can be calculated using IPD or summary data (sample sizes, SDs)<br>Assumes normality |
| ***Examining the relationship between mean and variance across the two arms*** |  |  |
| Difference in coefficient of variation (CVD) (26) | The difference in CoVs and its standard error are used to calculate a test statistic, whose square has approximate chi-squared distribution (1 degree of freedom) | Can be calculated using IPD or summary data (sample sizes, SDs, means)<br>Assumes normality<br>Data must be on a ratio scale with a meaningful zero<br>This test performs best if each $N_i > 10$ and each $\text{CoV}_i > 0.33$ (26). |
| Log of the ratio of coefficients of variation (logCVR) (25) | The log of the ratio of CoVs and the sampling variance are used to calculate a test statistic with approximate normal distribution, so a t-test is used to compare arms. | Can be calculated using IPD or summary data (sample sizes, SDs, means)<br>Can be made suitable for non-normal data by additions to the equation for sample variance<br>Data must be on a ratio scale with a meaningful zero |

<sup>a</sup>Note that this is called log of the variability ratio, logVR in these two references.

### 2.2 Examining the relationship between mean and variation across the two arms

If the mean is related to the variance for an outcome, then a homogenous treatment effect could lead to a difference in variance between the two arms of the trial. The CoV is the ratio of the SD to the mean: comparing CoVs between two arms will identify whether the standard deviation differs more, or less, between the two than would be predicted by the difference in means.

We describe two methods using CoV: a difference in CoVs (CVD, ^28^) and the log of the ratio of CoVs (logCVR, ^27^), Table 1 and eAppendix §2.

CoV should only be used when the outcome data are on the ratio scale, i.e. the scale has a clear definition of 0 and the ratio of two values has a meaningful interpretation. The CoV assumes that the SD is directly proportional to the mean. Therefore, it is only relevant for variables for which a sample mean of zero would imply a sample SD of zero. For example, a variable for which CoV would be appropriate would be serum cholesterol. This is measured on the ratio scale (a value of 6 is twice a value of 3), and there is a meaningful zero (the value 0 mg/dL indicates that there is no measurable cholesterol in 1 decilitre of blood). If a sample has a mean serum cholesterol of zero, this would indicate that all the values must be zero (as serum cholesterol cannot be negative), and therefore that the sample SD must be zero. We note that CoV has been used with outcomes which do not satisfy these criteria^17,20^.

### 2.3 Comparison of methods

The LMNCV method and Glejser’s test can incorporate covariates (which may be continuous or categorical), to examine whether the heterogeneity in outcome between the arms of the trial is explained by the covariates. The LMNCV method, Glejser’s, Levene’s and Bartlett’s tests can be defined for multiple *(k>* 2) arms. Bartlett’s test, VD, RoV, logSDR, CVD and logCVR can be calculated using only standard summary data (sample sizes, means and SDs).

All tests except Levene’s assume data are normally distributed: if data are normally distributed Levene’s test would be expected to have lower power. All the other tests are sensitive to non-normality of the outcome, for example if the subgroups have caused a bi-modal distribution or differing responses have caused skew. We note that normality usually cannot be verified when only summary data are available (although evidence against normailty, e.g. asymmetry of distributions, may be available by comparing mean and median).

### 2.4 Methods for use with summary data from meta-analyses

The approach to meta-analysis will depend on whether the result obtained from each trial is a statistical test or an estimate. In general, we favour estimation, preferring estimates of VD, RoV, logSDR and comparisons of CoVs (CVD and logCVR). Estimates that are accompanied by SEs can readily be meta-analysed using standard methods (here, the VD and CVD methods). RoV, logSDR and logCVR can be meta-analysed using bespoke methods using a random effects model with restricted maximum likelihood estimates (REML) of the ratios (RoV ^21^; logSDR and logCVR ^9,27^). We note that if variances within arms are very different across trials in a meta-analysis, ratio methods may be preferable.

Although not covered here, synthesis of findings from statistical tests from individual trials (e.g. Bartlett’s test and the F-test based on RoVs) could be undertaken using meta-analysis of p-values as described, for example, by Becker ^29^. These produce a global p-value to test the null hypothesis, although it can be difficult to determine whether failure to reject the null is due to small differences in variance or to an insufficient amount of evidence.

Previous analyses have implied that CoV should only be explored in a meta-analysis if the SDs and means within each trial arm are correlated ^9,17^. However, by simulating trial data (eAppendix §3), with (A) same CoV and (B) different CoV in the arms, we have shown that the correlation of the mean and SD from individual trials is not necessarily indicative of the CoV or whether the CoV differs between arms of the trial (eFigures 1&2). Thus, CoV should be used if (and only if) the outcome is a ratio variable with a true zero, irrespective of the observed correlation between SDs and means within trial arms.

## 3. Applied examples

### 3.1 Analysis of a single trial

We first apply the methods to individual participant data from a trial of therapist-delivered internet psychotherapy for depression in primary care ^30^. This RCT randomised 297 individuals to either usual care while on a waiting list for CBT (control) or usual care in addition to online CBT delivered by a therapist (intervention) ^30^. Baseline depression was measured using the Beck Depression Inventory (BDI) ^31,32^; individuals recruited to the trial had to have a BDI score of 14 or more. BDI is a self-report questionnaire with 21 statements that patients rank from 0-3 (i.e. total scores are integer and in the range 0-63), with a higher score indicating more severe depression ^31,32^. We investigated BDI at 4 months as a quantitative outcome. The equality of variances between the control and intervention arms was tested using: (1) LMNCV method (with and without adjusting for covariates); (2) Glejser’s test (with and without adjusting for covariates); (3) Levene’s test (using deviation from the mean, median and trimmed mean); (4) Bartlett’s test; (5) ratio of variances (F-test) method; and (6) logSDR method. The CVD and logCVR methods were not included as BDI is not ratio-scaled and therefore CoV is not a meaningful measure.

In order to examine the impact of differential dropout, the equality of variances between the control and intervention arms at baseline was also tested for (a) everyone; and (b) the subset of those remaining after excluding individuals lost to follow-up at 4 months, using Bartlett’s test, Levene’s test and the F-test.

### 3.2 Meta-analyses

We apply the summary data methods to two meta-analyses. The first summarises RCTs of computer-based psychological treatments for depression ^33^, including the single trial we assess above. Summary data were presented from 19 RCTs with intervention and control arms for 33 post-treatment effects. The outcomes were self-reported measures of depression, including BDI. As the measures of depression varied across trials, and to avoid double counting participants, we selected only those trials which measured BDI (or derivatives of BDI) and kept only one post-treatment effect per trial. As the subset selected were all measuring BDI, we could meta-analyse the VD, RoV and logSDR across trials. However, we did not include the CVD or logCVR methods as BDI is not ratio-scaled.

Our second example is a Cochrane Review examining HMG CoA reductase inhibitors (statins) for people with chronic kidney disease ^34^. We chose this example because there is evidence that some people may respond to statins better than others ^35^. The data presented are from analysis 1.14 in the review, for 22 trials reporting the effect of statins versus placebo or no treatment on LDL cholesterol, reported in mg/dL. LDL cholesterol is measured on a ratio scale, with a meaningful zero, and thus we meta-analysed the VD, RoV, logSDR, CVD and logCVR across trials.

## 4. Results

### 4.1 Analysis of a single trial

Of the 297 individuals recruited to the trial at baseline, 210 completed 4-month follow up (113 in the intervention arm and 97 in the control arm, Table 2) ^30^. The BDI score had decreased in both arms, with a larger magnitude decrease in the intervention arm. The BDI scores were normally distributed at baseline, but not at the 4-month follow up (eFigure 3).

**Table 2:** The baseline Beck Depression Inventory (BDI) score and outcome BDI score at 4 months from the trial described in Kessler 2009.

| Timepoint | Group 1 (Intervention) |  |  | Group 2 (Control) |  |  |
| --- | --- | --- | --- | --- | --- | --- |
|  | N | Mean | SD | N | Mean | SD |
| Baseline | 149 | 32.8 | 8.3 | 148 | 33.5 | 9.3 |
| 4 months | 113 | 14.5 | 11.2 | 97 | 22.0 | 13.5 |

Table 3 shows the results of all tests on the variance of BDI at 4 months. Even though the data at 4 months were not normally distributed, the conclusions from all tests were similar to Levene’s test, with the p-values for all but adjusted model 1 being between 0.03 and 0.07, giving weak evidence of lower variance in the intervention arm of the trial. Including baseline BDI score (adjusted model 2 in the LMNCV method and the Glejser test) largely removed any evidence of difference in variance between the arms (p>0.2). This implies the effect of the intervention would be the same for all individuals with the same baseline BDI score.

**Table 3:**
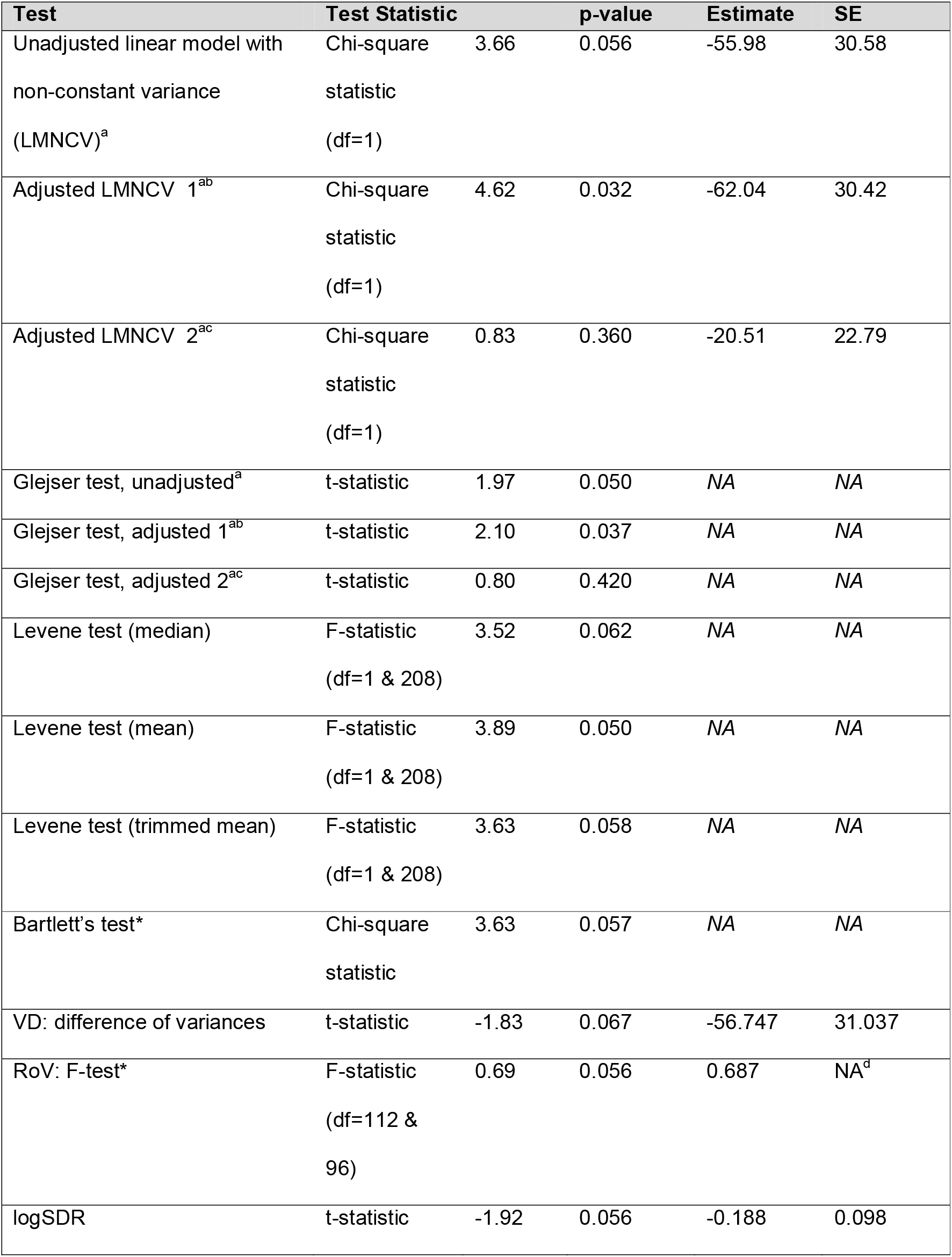
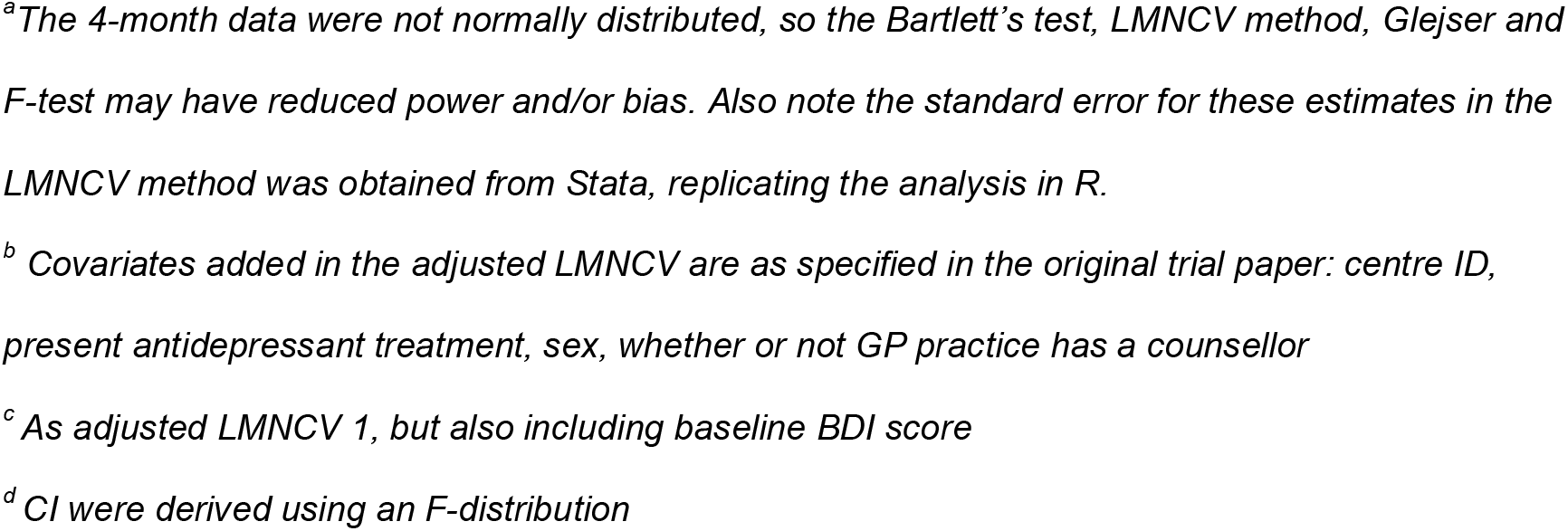
Tests for difference in variance in BDI score at 4 months, between the intervention and control arms from the single trial exploring the effect of a CBT intervention on depression (28).

| Test | Test Statistic |  | p-value | Estimate | SE |
| --- | --- | --- | --- | --- | --- |
| Unadjusted linear model with non-constant variance (LMNCV)<sup>a</sup> | Chi-square statistic (df=1) | 3.66 | 0.056 | -55.98 | 30.58 |
| Adjusted LMNCV 1<sup>ab</sup> | Chi-square statistic (df=1) | 4.62 | 0.032 | -62.04 | 30.42 |
| Adjusted LMNCV 2<sup>ac</sup> | Chi-square statistic (df=1) | 0.83 | 0.360 | -20.51 | 22.79 |
| Glejser test, unadjusted<sup>a</sup> | t-statistic | 1.97 | 0.050 | NA | NA |
| Glejser test, adjusted 1<sup>ab</sup> | t-statistic | 2.10 | 0.037 | NA | NA |
| Glejser test, adjusted 2<sup>ac</sup> | t-statistic | 0.80 | 0.420 | NA | NA |
| Levene test (median) | F-statistic (df=1 & 208) | 3.52 | 0.062 | NA | NA |
| Levene test (mean) | F-statistic (df=1 & 208) | 3.89 | 0.050 | NA | NA |
| Levene test (trimmed mean) | F-statistic (df=1 & 208) | 3.63 | 0.058 | NA | NA |
| Bartlett's test* | Chi-square statistic | 3.63 | 0.057 | NA | NA |
| VD: difference of variances | t-statistic | -1.83 | 0.067 | -56.747 | 31.037 |
| RoV: F-test* | F-statistic (df=112 & 96) | 0.69 | 0.056 | 0.687 | NA<sup>d</sup> |
| logSDR | t-statistic | -1.92 | 0.056 | -0.188 | 0.098 |

<sup>a</sup>The 4-month data were not normally distributed, so the Bartlett's test, LMNCV method, Glejser and F-test may have reduced power and/or bias. Also note the standard error for these estimates in the LMNCV method was obtained from Stata, replicating the analysis in R.
<sup>b</sup> Covariates added in the adjusted LMNCV are as specified in the original trial paper: centre ID, present antidepressant treatment, sex, whether or not GP practice has a counsellor
<sup>c</sup> As adjusted LMNCV 1, but also including baseline BDI score
<sup>d</sup> CI were derived using an F-distribution

The analysis of baseline variances showed no differences between the two arms at baseline, even when restricting to only those with follow up data at 4 months (eAppendix §4.2, eTable 4).

### 4.2 Meta-analyses

Our simulations confirmed that power to detect heterogeneity in single trials was low unless the trial was very large (see eAppendix §6). Therefore, we next examined the methods within a meta-analysis setting.

Restricting the meta-analysis on computer-based psychological treatments for depression ^33^ to trials reporting BDI score gave a subset of 11 trials, varying in size from 44 to 216 participants. Two of the 11 trials showed evidence of greater variance in the control arm using RoV (eTable 5, Figure 1). One of these also had evidence of greater variance in the control arm using the VD. The meta-analysis gave evidence of greater variance in the control arm (RoV 0.82 [95% CI: 0.67, 1.00]; VD fixed-effect estimate −19.13 [95% CI: −32.79, −5.48], random-effects mean −18.19 [95% CI: −33.80, −2.58]). Using logSDR gave the same trends as the RoV test, eTable 5.

**Figure 1:**
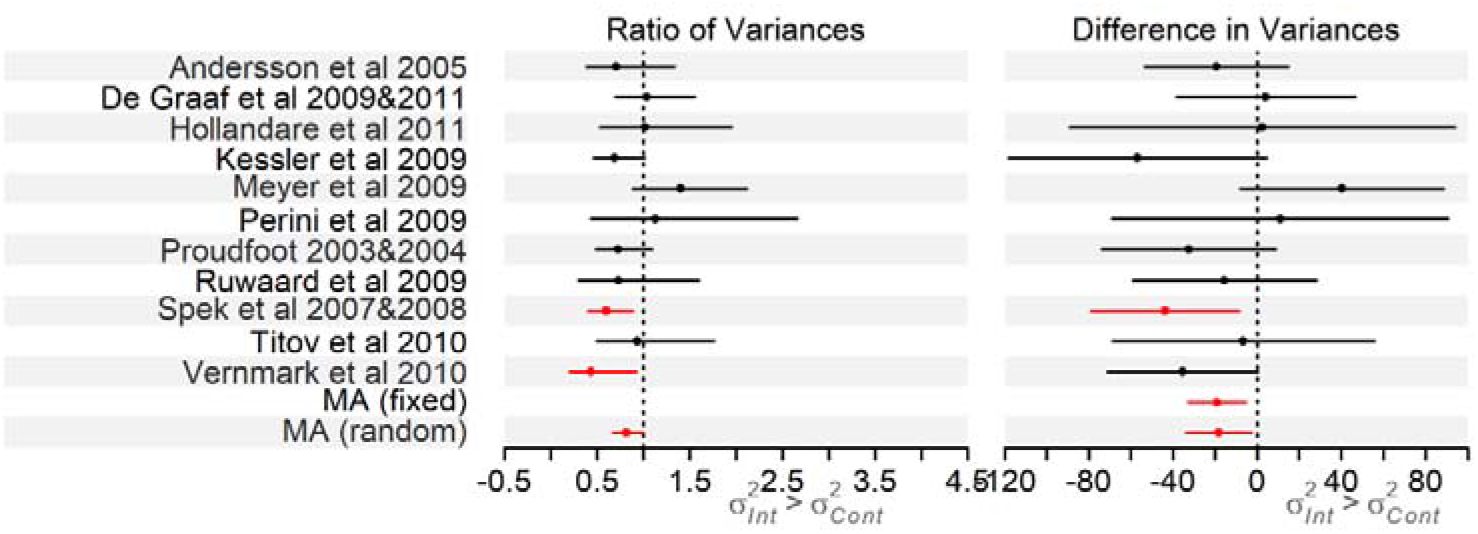
Forest plot of the RoV and VD analyses for the trials in the Richards et al meta-analysis on computer-based psychological treatments for depression ^33^, results in eTable 5 (note we do not plot the results of the logSDR analysis as trends are the same as the RoV analysis).

The 22 trials in the meta-analysis reporting the effect of statins versus placebo or no treatment on LDL cholesterol ^34^, varied in size from 199 to 374 total participants. Two of the trials showed evidence of greater variance in the control arms using the VD (eTable 6, Figure 2). Using RoV, five trials had evidence for greater variance in the control arm (RoV<1) and for one trial there was evidence of greater variance in the intervention arm (RoV>1). logSDR gave the same trends as the RoV, eTable 6. There is evidence that the CoV was greater in the intervention arm in four trials and greater in the control arm in one trial (Figure 2): the same trends were identified with a test of logCVR (eTable 6).

**Figure 2:**
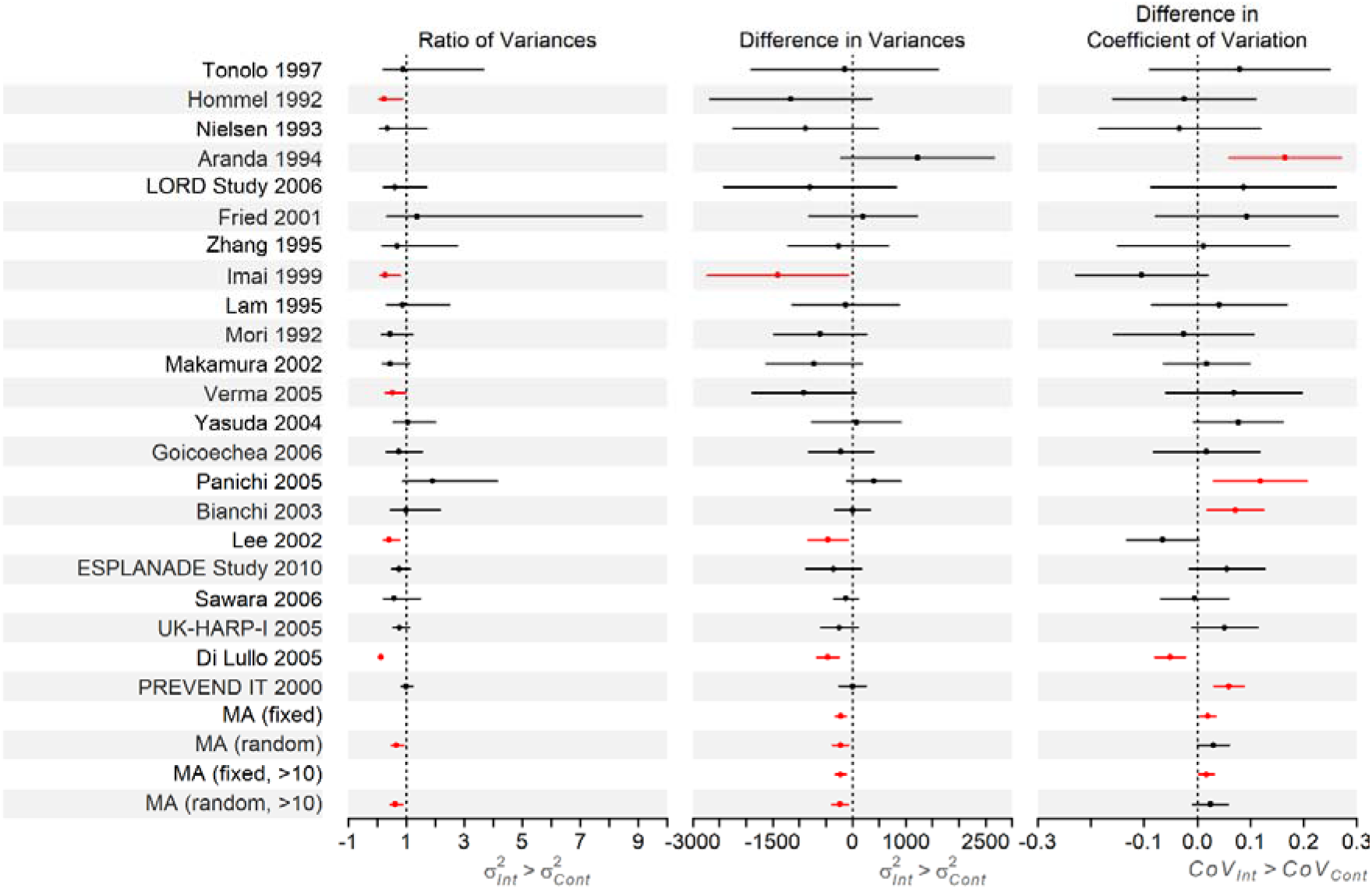
Forest plot of the RoV, VD and CVD analyses of the trials in the Palmer et al meta-analysis reporting the effect of statins versus placebo or no treatment on LDL cholesterol ^34^, results in eTable 6. We have not plotted the RoV results for Aranda 1994 as the RoV for this trial is on a much larger scale than the others (RoV=9.51 [95% CI: 1.90, 47.49]), however it is included in the overall analysis. Note we do not plot the results of the logSDR or logCVR analyses as trends were the same as the RoV and CVD analyses, respectively.

The meta-analysis of the VD gave evidence of greater variance in LDL cholesterol in the control arm (fixed-effect estimate −220.36 [95% CI: −318.84, −121.87] mg^2^/dL^2^, random-effects mean −226.33 [95% CI: −376.77, −75.90] mg^2^/dL^2^), which remained when only trials with more than 10 cases in both arms were included (excluding 6 trials, fixed-effect −223.51 [95% CI: −323.90, −123.12] mg^2^/dL^2^, random-effects −233.17 [95% CI: −388.82, −77.53] mg^2^/dL^2^). The pooled RoV also showed evidence of greater variance in the control arm 0.66 [95% CI: 0.48, 0.91] (eTable 6, Figure 2). The ratio was further from the null (0.62 [95% CI: 0.44, 0.87]) if the six smallest trials were excluded. However, there was weak evidence of a difference in CoV between arms (difference in CoV for intervention compared to control arm of 0.02 [95% CI: 0.01, 0.03] for fixed effects, and 0.03 [95% CI: −0.00, 0.06] from a random-effects model; and with the 6 smallest trials excluded: fixed = 0.02 [95% CI: 0.00, 0.03], random = 0.03 [95% CI: −0.01, 0.06]). This indicates that the CoV is larger in the intervention arm than in the control arm, i.e. the SD is a larger multiple of the mean in the intervention than the control arm. This suggests weak evidence of more variation in the intervention arm than would be expected given the difference in means, which could be due to statins having a greater effect for some people than others.

## 5. Discussion

We have presented methods for examining differences in outcome variance between the two arms in an RCT, in order to identify between-individual heterogeneity of effects of the intervention. We have added to existing methods by: showing how to use regression-based methods to examine the effects of covariates on variation, where individual participant data are available; applying a difference of variances test to summary data in meta-analyses, alongside the RoV, logSDR and logCVR methods already existing; and noted when the CoV test is not appropriate. We suggest that CoV methods, which explore whether the difference in variance is due to a difference in means, are only used where the outcome has a meaningful zero and is on a ratio scale.

Differences in variance could be caused by many factors. One is the existence of patient characteristics that influence the effectiveness of the intervention (effect modifiers), which could manifest as subgroups between which the intervention (or control) treatments have different effects ^9^. For example, the intervention may have a different effect in those with worse (or better) values at baseline, or outcomes in the control arm may vary due to differences in “usual practice”. If there are differences in variance, further studies may be needed to find the effect modifiers that define the subgroups.

Other potential explanations for differences in variance between arms of a trial are: differential dropout between arms of the trial, where dropout is related to outcome; non-compliance with the intervention; subgroups that are differently engaged with the intervention (for example, therapist effects) or an intervention that impacts on within-person variability ^9^. Investigation of other factors relating to variation would require individual or stratified summary data on these factors – such as pre-treatment severity, or marital status moderating the response to CBT ^36^. Another explanation for differences in variance is model misspecification (for example if the errors follow a non-normal distribution, or if the errors are not independently distributed). Investigation of misspecification of the model would require IPD, and examination of the model within each trial.

Simulations confirmed that power to detect heterogeneity in single trials was low unless the trial was very large ^2^. RCTs would need to increase their sample size by orders of magnitude to be powered to detect difference in variance and allow further analysis. This might be prohibitively expensive in time and money, and it may not even be feasible to recruit enough individuals with the required condition to the trial ^37^. In this case, as large a sample size as possible is appropriate, and improved reporting, for example, giving detailed summary data across both trial arms, would allow a trial to be included in a meta-analysis using methods we have described here.

Smaller variance in the intervention than the control arm was observed in both meta-analyses presented here, but without IPD it was not possible to explore this further. With IPD, the factors associated with the variance can be examined directly, as in the single trial example presented here ^30^. These factors might be used to predict the effect of the intervention in external populations or applied in personalised medicine. The slightly lower variance in the intervention arm in the single trial ^30^ and meta-analysis of effects of CBT in depression ^33^ may also be partly because BDI is bounded at 0 and floor (or ceiling) effects can reduce variance.

Another possible cause of differences in variance between two arms of a trial is that the variance is related to the mean, and the intervention causes a mean difference in the outcome. This is clearly shown in our second meta-analysis example, examining the effect of statins on LDL cholesterol ^34^. There was evidence that the variance of the outcome was lower in the intervention than the control arm. As the intervention lowered mean cholesterol levels, this implied that statins had a greater effect on those with initially higher cholesterol levels. However, the CoV results indicated that this difference in means was associated with the difference in variation. This led to the important conclusion that the variance in the intervention arm was actually a little larger than would have been expected, given the difference in means – thus providing some (weak) evidence that there was heterogeneity in the effect of statins on LDL cholesterol.

It is important to use the right method for the data. If IPD were available, Levene’s and Glejser’s tests could also be used, and comparing results across tests would explore the impact of any non-normality of the data. For meta-analysis of individual trials, the assumption of normality should be checked as far as possible (e.g. by using data presented within each paper such as mean, median, SD). Ratios are appropriate where different scales are used across different trials or where the same scale is used but the mean is very different, as in these situations a difference in variances test may not be appropriate. These methods may be biased when the arms are not independent (for example in cross-over trials) ^14^.

These methods for quantifying variance between treatment arms are applicable not just to RCTs, but also to differences in variance of continuous outcomes according to genotype in genetic epidemiological studies ^38-40^. Differences by genotype can be considered as analogous to differences by treatment arm in an RCT ^41,42^, indeed the progenitor of RCTs, RA Fisher, considered the factorial nature of Mendelian inheritance as the model for randomization in experiments ^43-45^. In this regard, difference in variance by allele count at, e.g., a single-nucleotide polymorphism (SNP) locus, is taken as evidence of the presence of either epistasis or gene-environment interaction ^38-40^. A second potential application is within Mendelian randomization (MR) implemented within an instrumental variables (IV) analysis framework ^46,47^. An interpretative issue relates to the assumption of homogeneity of the effect of the instrument on the exposure, since violations of this would suggest that IV effect estimates may not apply to the entire study sample. Indeed the exposure under investigation may have effects in opposite directions among different members of the study sample. The assumption of such homogeneity is sometimes refered to as the 4^th^ IV assumption ^48^, for which there are various weaker versions (including monotonicity of the instrument-exposure association ^48^). As non-homogeneity in the genetic variant – exposure association would lead to non-homogeneity in the genetic variant – outcome assocation, then as long as either the exposure or outcome allow variance estimation then an umbrella test of presence and degree of violation of IV4 is possible. This approach would, of course, apply to IV analysis in general and not just when this is within an MR context.

Whilst conclusions from randomised trials are usually expressed in terms of average effects of an intervention, individuals will want to know how well they personally will respond to an intervention. Grouping subjects according to an observed response is open to bias ^49^. An alternative way to examine variation in response, without having to specify and measure effect modifiers, is to examine differences in variability between the trial arms. We have described different ways of doing this with IPD or using summary data. Given the low power to explore heterogeneity of variance in individual trials, we suggest that meta-analyses should be used where possible. It is important to test the coefficient of variation between trial arms, and also to consider the other explanations (e.g. compliance) for heterogeneity of variance: using multiple different approaches can help explore these possibilities.

## Data Availability

The data for the single RCT example used in this paper are not available with this article and requests should go via David Kessler lead author of the original trial paper. Data for the meta-analysis examples are provided in the supplementary material.
Code for each method in R is provided online at the link below.

https://github.com/harrietlmills/DetectingDifferencesInVariance

## Acknowledgements

We thank Luke Prendergast for providing example code based on his 2016 paper “Metaanalysis of ratios of sample variances”.

